# Impact of continuous labour companion- who is the best: A comprehensive meta-analysis on familiarity, training, temporal association, and geographical location

**DOI:** 10.1101/2024.02.02.24302191

**Authors:** DMCS Jayasundara, IA Jayawardane, SDS Weliange, TDKM Jayasingha, TMSSB Madugalle

**Affiliations:** Department of Obstetrics and Gynaecology, Faculty of Medicine, University of Colombo, Colombo, Sri Lanka; De Soysa Maternity Hospital, Colombo, Sri Lanka; Department of Community Medicine, Faculty of Medicine, University of Colombo, Colombo, Sri Lanka; National Hospital, Kandy, Sri Lanka

**Keywords:** Continuous labour companion, Meta-analysis, Trained companion, Untrained companion, Familiar companion, Unfamiliar companion, Temporal association, Asia, Africa, Europe

## Abstract

**Background:** Continuous labour support is widely acknowledged for potentially enhancing maternal and neonatal outcomes and smoothing the labour process. However, existing literature lacks a comprehensive analysis of the optimal characteristics of labour companions, particularly in comparing the effects of trained versus untrained and familiar versus unfamiliar labour companions across diverse geographical regions and pre and post-millennial. This meta-analysis addresses these research gaps by providing insights into the most influential aspects of continuous labour support.

**Methodology:** A thorough search of PubMed, Google Scholar, Science Direct, International Clinical Trials Registry Platform (ICTRP), ClinicalTrials.gov, Research4Life, and Cochrane Library was conducted. Study selection utilised the semi-automated tool Rayyan. The Cochrane risk-of-bias (RoB2) tool and funnel plots gauged the risk of bias. Statistical analysis employed RevMan 5.4, using Mantel-Haenszel statistics and random effects models to calculate risk ratios with 95% confidence intervals. Subgroup analyses were performed for different characteristics, including familiarity, training, temporal associations, and geographical locations. The study was registered in INPLASY. (Registration number: INPLASY202410003)

**Results:** Thirty-five randomised controlled trials (RCTs) were identified from 5,346 studies. The meta-analysis highlighted significant positive effects of continuous labour support across various outcomes. There was a substantial improvement in the 5-minute APGAR score < 7, with an effect size of 1.52 (95% CI 1.05, 2.20). Familiar labour companions showed a higher effect size in reducing tocophobia, 1.73 (95% CI 1.49, 2.42), compared to unfamiliar companions, 1.34 (95% CI 1.14, 1.58). Differences were noted between trained and untrained companions, favouring untrained companions in reducing tocophobia and the cesarean section rate. Studies conducted after 2000 had a more significant impact on decreasing labour duration. Geographical variations indicated more pronounced effects in Asia and Africa than in Europe.

**Discussion and Conclusion:** The meta-analysis underscores the benefits of labour companionship, particularly in facilitating the parturient experience of spontaneous labour. The impact is more pronounced in specific subgroups, such as familiar companions, untrained companions, recent studies, and studies conducted in Asia and Africa. The study recommends integrating labour companionship into obstetric care pending further research, standardisation, and awareness initiatives to enhance maternal and neonatal outcomes. Challenges such as study heterogeneity, insufficient data on companion training, and temporal outcome variations are acknowledged.

## Introduction

The emotional process of labour and childbirth is often a fearful and stressful event for a pregnant mother (1). In most cultures, the tradition of supporting a woman in labour is a community event with multiple participants other than the designated healthcare provider. The fear and anxiety of childbirth are augmented by an unfamiliar hospital environment, medical jargon, procedures, interventions and transient separation from the family during labour (2). The woman feels a sense of loss of control, isolation and fear, peaking the level of anxiety (2). To cope with this tocophobia, pregnant women sometimes choose cesarean section over natural birth (3). Increased anxiety makes the woman more vulnerable to increased pain perception, prolonging the duration of labour and contributing to dystocia (4). The pain and anxiety during labour increase the endogenous catecholamine release, causing ineffective uterine contractions and decreased placental blood flow (5). An inefficient labour process may cause fetal and maternal complications, including the risk of fetal or neonatal hypoxia and death, infection, physical damage in the newborn, postpartum haemorrhage, maternal infection and psychological distress due to anxiety, lack of sleep and fatigue (4).

Different clinical settings have adopted strategies to alleviate tocophobia, facilitating a smooth labour process. Support methods include having an accompanying companion for continuous labour support, induced sleep, hydrotherapy, and the Lamaze relaxation method (6). The labour companion can be a non-caregiving nurse, midwife, friend, relative, family member, husband or a person trained in supporting labour (doula) (3). WHO defines labour support as the supportive care provided to women during labour, including emotional support, physical comfort, advice and information giving (5). WHO also recommends that a parturient should have a birth companion of her choice. However, it is not practised in many developing countries (7).

Having a companion for continuous labour support facilitates a smooth labour process, improving the maternal psychological status and fetal/neonatal well-being. Reported advantages include an increase in spontaneous vaginal births, reduced demand for analgesics, reduced need for oxytocin for labour augmentation, shorter duration of labour, decreased need for cesarean sections, minimal perineal trauma, and reduced requirement for instrumentation during labour, facilitating a smooth labour process (8–11). Maternal psychological well-being is improved by lowering tocophobia, reduced postpartum depression and anxiety, and improved self-esteem and satisfaction measured postpartum (2,12,13). Fetal/neonatal well-being is enhanced by the early establishment of exclusive breastfeeding, early skin-to-skin contact, reduced neonatal hospital stay, and the need for neonatal resuscitation (14,15).

The quality of labour support and its beneficial outcomes depend on the type of companion used (16). The labour companion can be trained or untrained and familiar or unfamiliar to the parturient. The evidence regarding the “best labour companion” is controversial, and studies do not show a clear consensus.

The rates of severe tocophobia, measured similarly, vary in different countries, and the reasons are unknown (17). The prevalence of tokophobia was lower in the early years (1980s, 1990s) compared to more recent years (2000 onwards) (18). The beneficial effects of a labour companion can be more pronounced in some countries compared to others and may have changed over time. The present meta-analysis aims to describe the characteristics of the most effective labour companion, highlighting the differences in beneficial effects of having a labour companion among different geographical regions and timelines.

## Materials and methods

### Search strategy

PubMed, Science Direct, Cochrane Library, Google Scholar, ClinicalTrials.gov and International Clinical Trials Registry Platform (ICTRP) were searched on 04/07/2023 (the date of the most recent search). To identify relevant studies, a set search strings such as “Labour companion,” “Birth partner,” “Doula,” “Labour support person,” “Childbirth coach,” “Labour assistant,” “Labour coach,” “Birth attendant,” “Labour caregiver,” “Maternity support person,” “Childbirth companion,” “Labour ally,” “Labour chaperon,” “Pregnancy outcome,” “Obstetric outcome,” “Delivery outcome,” “Birth outcome,” “Fetal outcome,” “Newborn outcome,” “Infant outcome,” “Neonatal outcome”, and “Baby’s outcome” were employed, with Boolean expressions “AND” and “OR” used appropriately to construct precise search queries. Initially, the literature search was conducted without filters. Then, the results were refined using advanced search options like full-text articles and randomised controlled trials.

A manual search strategy was also applied to ensure inclusivity, focusing on identifying any missing studies by reviewing the most cited ten meta-analyses within the same databases.

The study selection process was carried out meticulously in two rounds using a semi-automated tool, Rayyan (19), with one author as the reviewer and another as a collaborator, employing a blind approach. In the first round, titles and abstracts were screened, eliminating duplicates and ineligible entries, with conflicts resolved by the reviewer. The second round involved a similar blind approach for full-text screening, again with conflicts resolved by the reviewer. The authors were contacted in cases requiring additional information. The study selection process was transparently reported using the PRISMA 2020 flow diagram for updated systematic reviews (20).

A detailed search strategy is given as a separate file under supporting information (S10 File). A protocol exists for the current study, and a copy of the protocol is given as supporting information (S11 File).

### Inclusion and exclusion criteria

Randomised controlled trials (RCTs) with full-text articles reporting results related to low-risk women with a single fetus in cephalic presentation, admitted during early labour (cervical dilation 3-4 cm) with no contraindications for vaginal delivery were included in the study. Studies reporting women with medical or psychiatric diseases, previous cesarean section, genital abnormalities, fetal distress and any fetal anomaly were excluded. Review articles, case reports, documents, or observational studies were excluded.

### Data extraction and quality assessment

Key study characteristics were extracted and organised into predefined tables for outcome measures concerning the facilitation of the labour process, maternal psychological well-being and fetal well-being. Concerned outcome measures were spontaneous vaginal birth, tocophobia, postpartum depression, admission to a special care nursery, exclusive breastfeeding, analgesic usage, synthetic oxytocin usage, duration of labour, LSCS rate, labour pain, instrumental vaginal delivery, perineal trauma, 5-min APGAR score, neonatal hospital stay, maternal anxiety, maternal self-esteem and maternal satisfaction. To ensure the integrity of the research, a second author independently reviewed the entire process, minimising the potential for bias. The quality of each RCT was assessed using the Cochrane risk-of-bias (RoB2) tool (21). Random sequence generation, allocation concealment, performance bias, detection bias, attrition bias, reporting bias, and other biases are used as criteria in RoB2. Funnel plots were employed to gauge publication bias, with any deviation from the expected funnel-shaped distribution as an indicator of potential publication bias.

### Statistical analysis

We used RevMan version 5.4 to analyse the following outcome measures reported by more than ten RCTs - spontaneous vaginal birth, tocophobia, use of analgesics, need for synthetic oxytocin, duration of labour, LSCS rate, instrumental vaginal delivery and 5-min APGAR score. The Mantel-Haenszel statistical method, random effects analysis model, and risk ratio with a 95% confidence interval (CI) as effect measures were used for dichotomous data. For the continuous data inverse variance statistical method, the random effects analysis model and standard mean difference as effect measures were used. We assessed heterogeneity with the I2 statistic, considering p value < 0.1 or I2 > 50% indicators of significant heterogeneity. Subgroup analyses were conducted to compare the effects of trained vs. untrained labour companions, familiar vs. unfamiliar labour companions, studies before vs. after 2000 and studies in different geographical locations.

## Results

### Search results, study characteristics and quality assessment

Figure 1 shows the PRISMA 2020 flow diagram for study selection. We identified 5346 studies from 7 databases and a manual search. After considering exclusion and inclusion criteria, 35 studies were selected for analysis.

**Figure 1.**
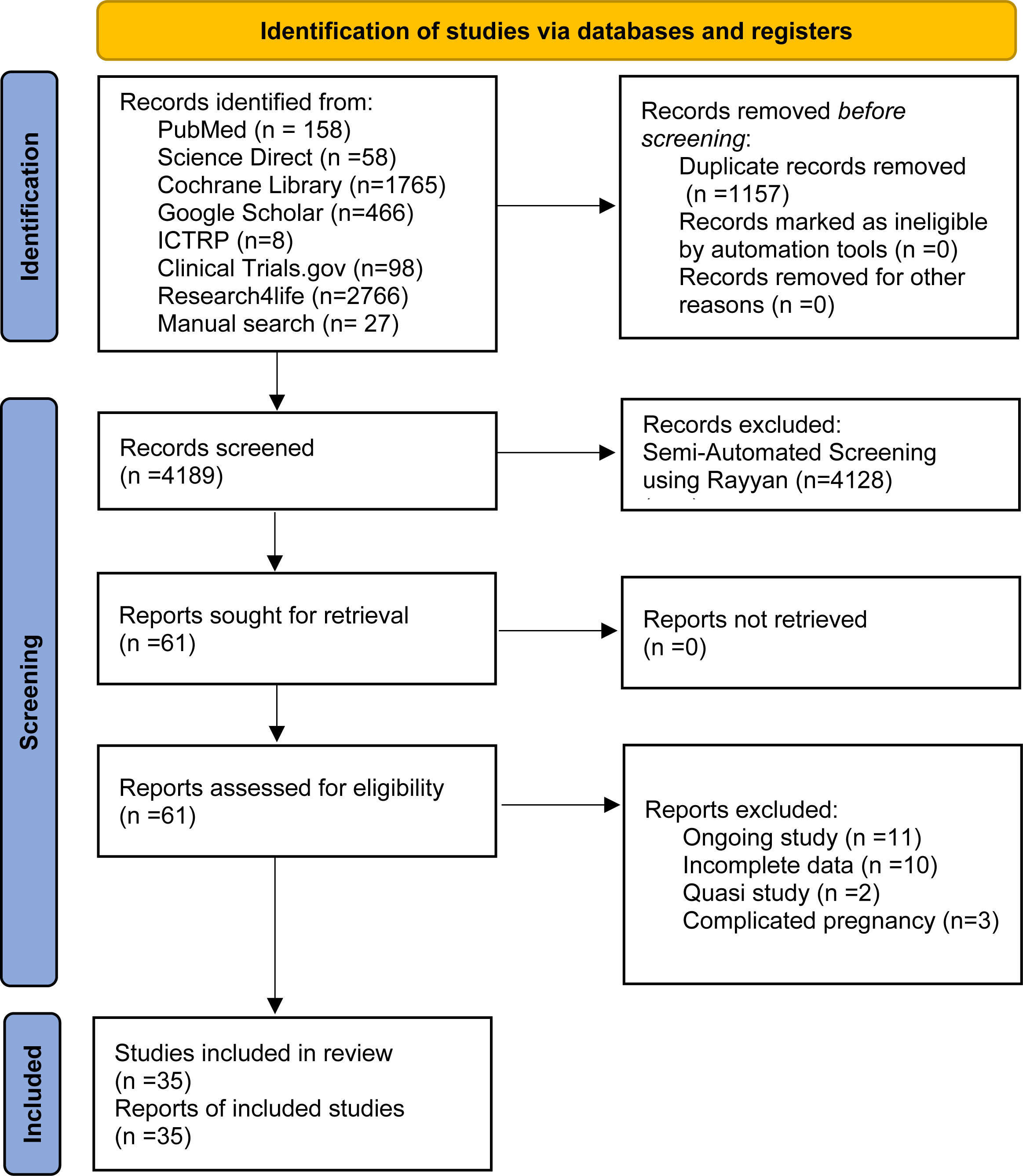
PRISMA 2020 flow diagram for study selection after initial filtering.

Table 1 summarises the key characteristics of 35 RCTs, including the year of the study, country, number of participants, a description of the type of labour companion, and outcome measures. Studies span from 1986 to 2022 from various geographical regions: Asia, Africa, Europe, North America, South America and Australia. Three studies (8.57%) have less than 100 participants, while 7 (20%) have more than 500 participants. Hodnett 2002 from the USA has the highest number of participants at 6915. Different studies have used labour companions with varying characteristics, such as familiar, unfamiliar, trained and untrained. Twenty-three studies (65.71%) used trained labour companions, while 20 (57.14%) used unfamiliar ones. The individual studies have examined different outcome measures, with a recent emphasis on maternal psychological well-being.

**Table 1.**
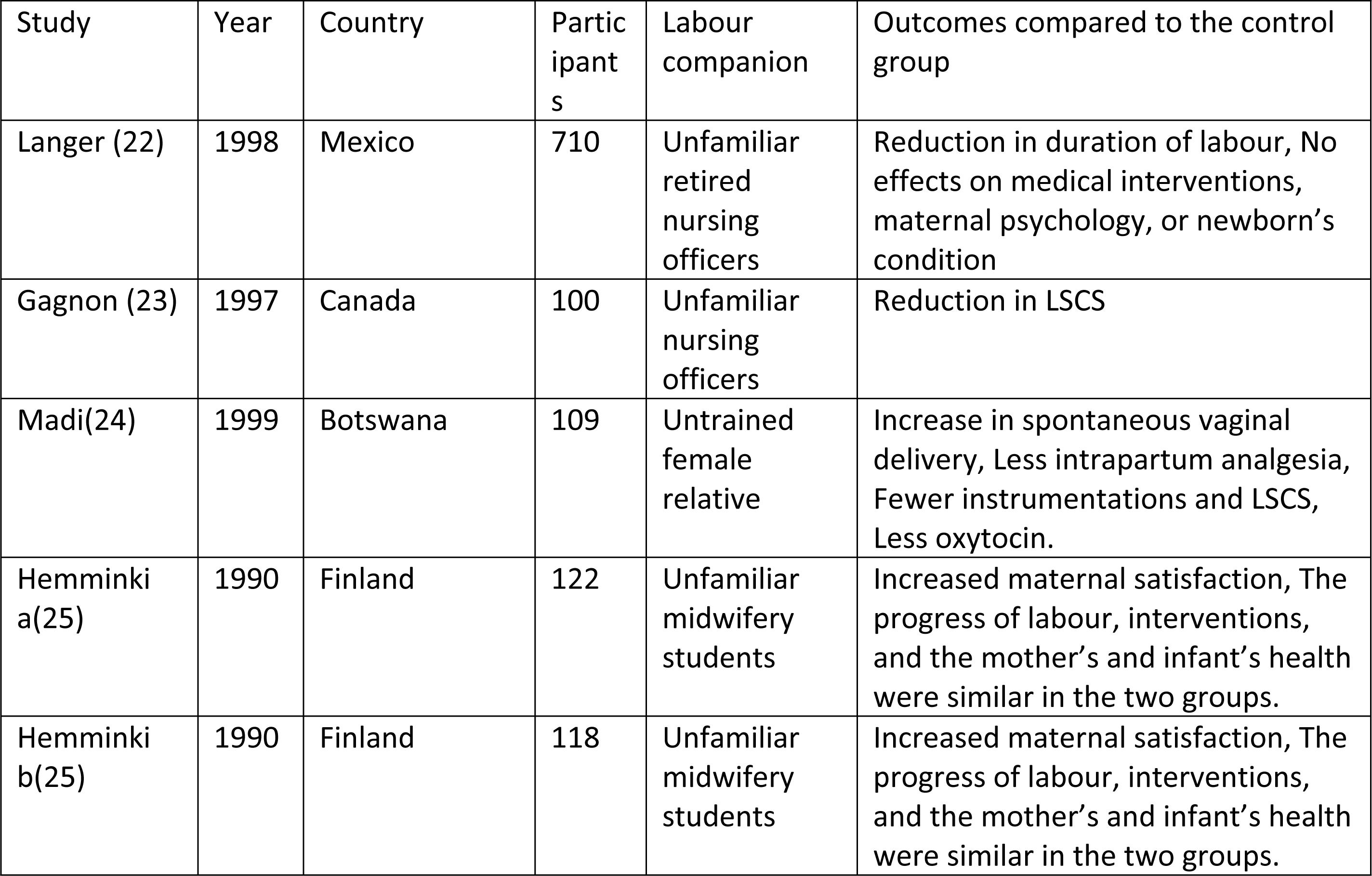
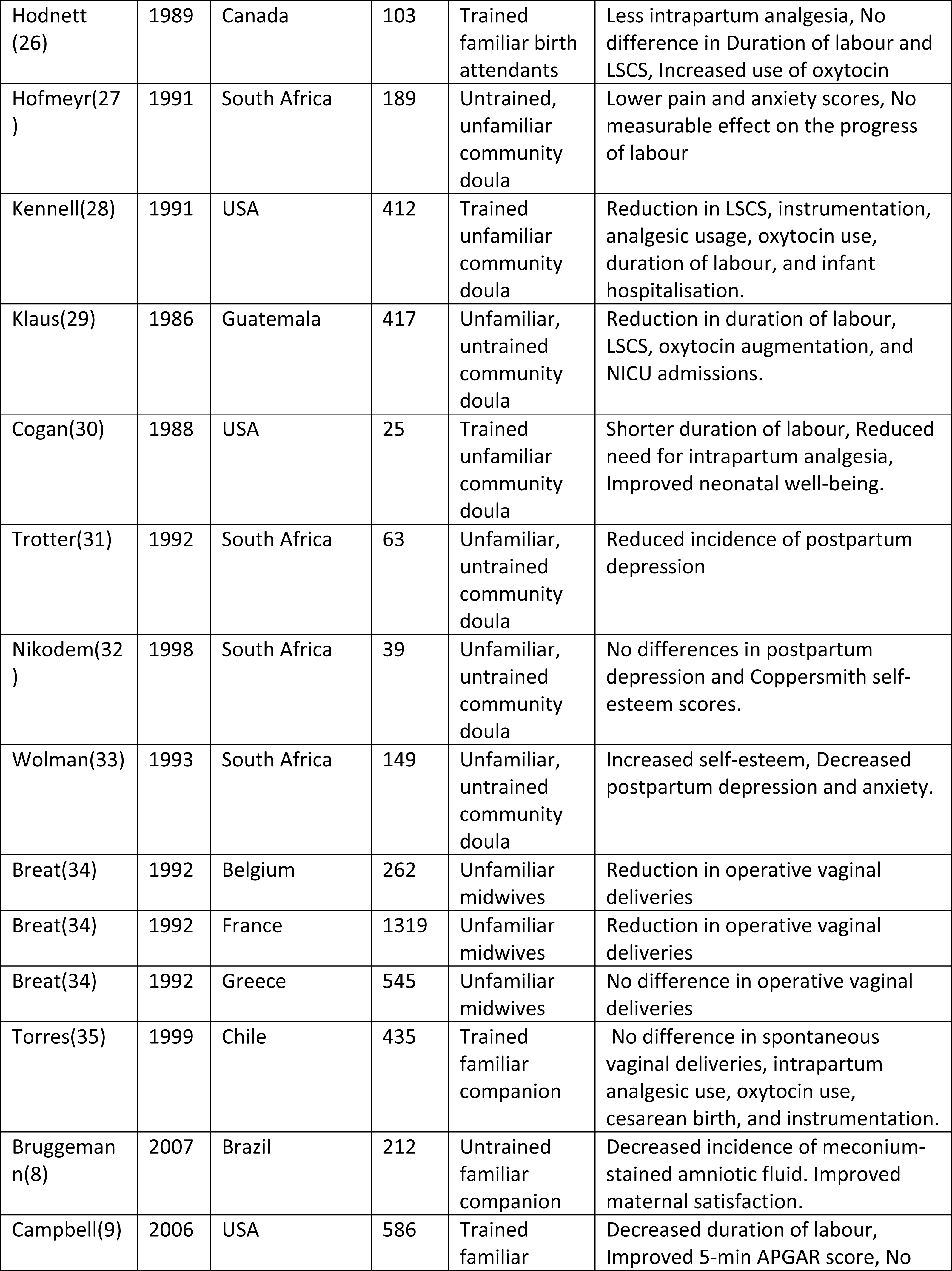
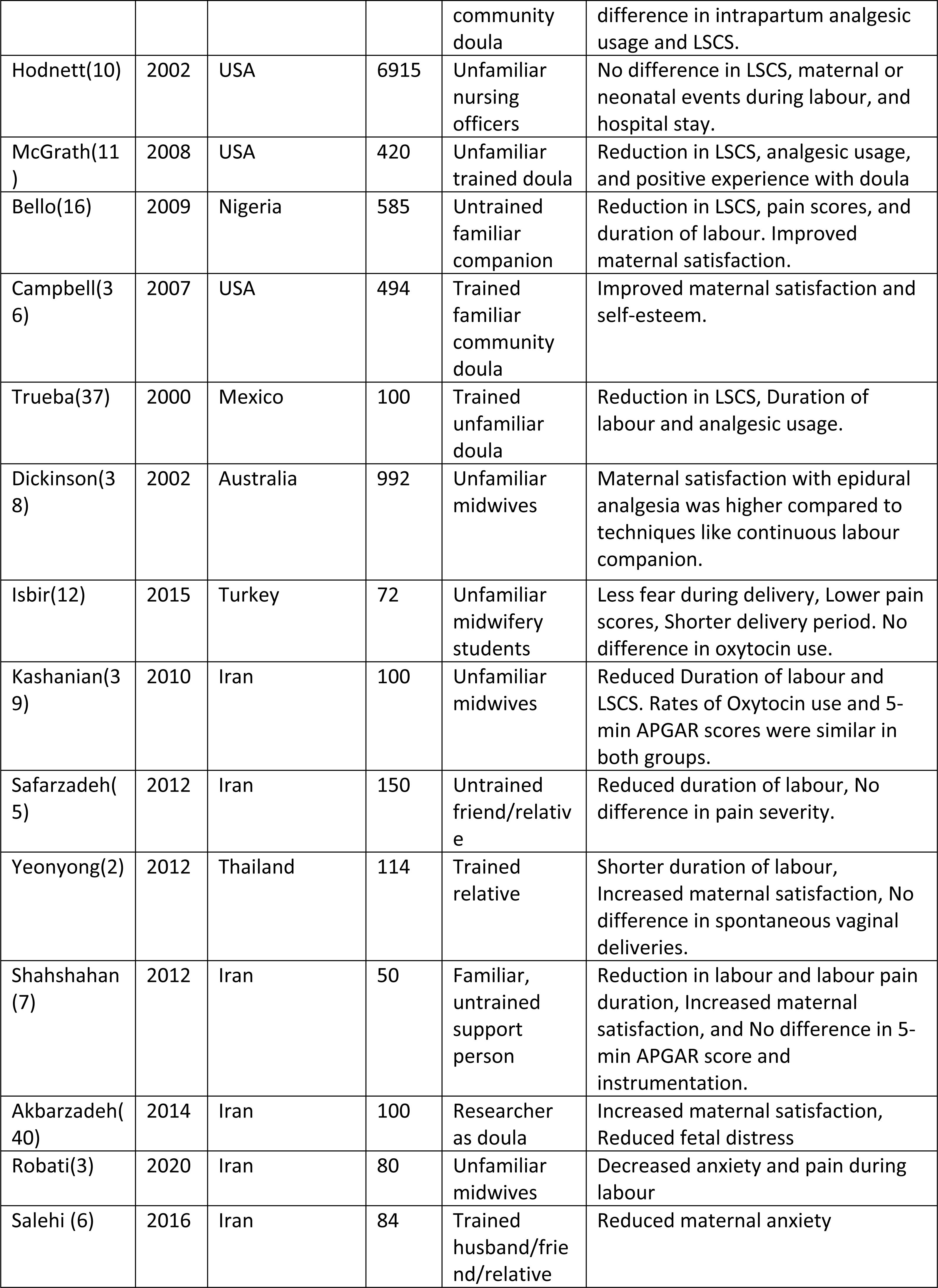
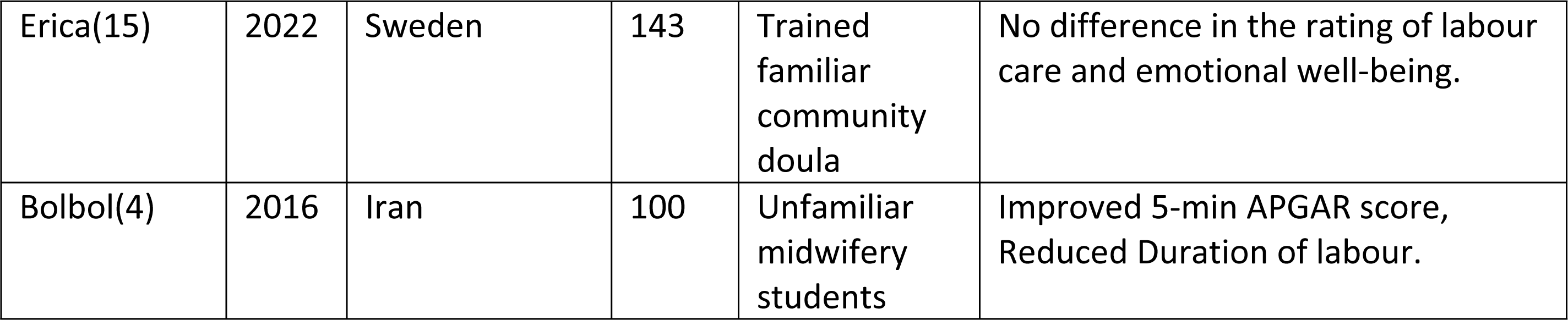
Key characteristics of RCTs analysed in the meta-analysis.

The funnel plots exhibited symmetry, suggesting minimal publication bias. Figures 2 and 3 summarise the RoB2 assessment of RCTs. All the studies show an overall high risk of bias due to having unclear risk for multiple domains or a high risk of bias in at least one domain (41). The highest risk of bias is reported in the blinding of participants and personnel (Figure 3).

**Figure 2.**
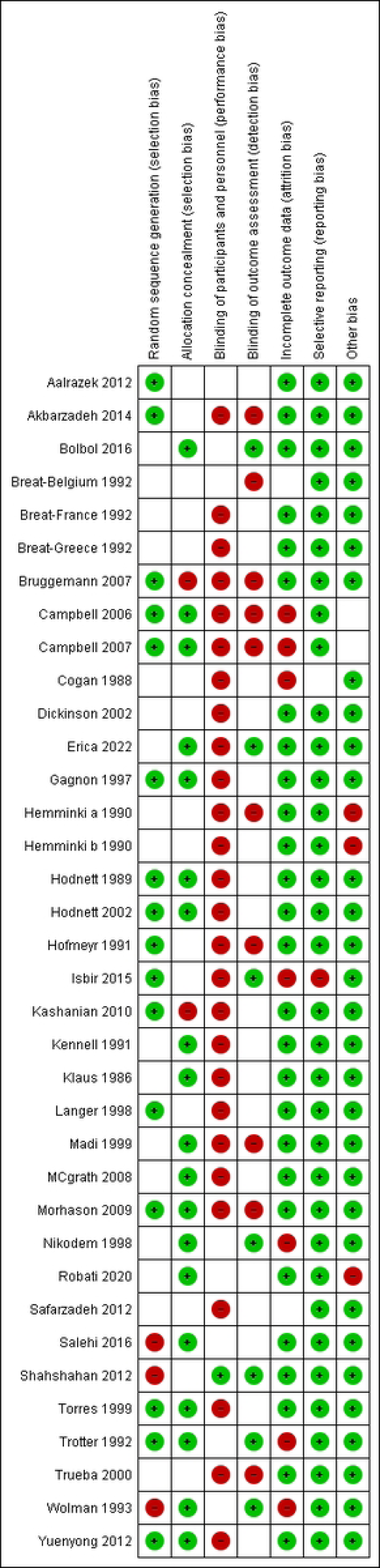
Risk of bias summary. Green: Low risk, Red: High risk, Blank: Unclear risk

**Figure 3.**
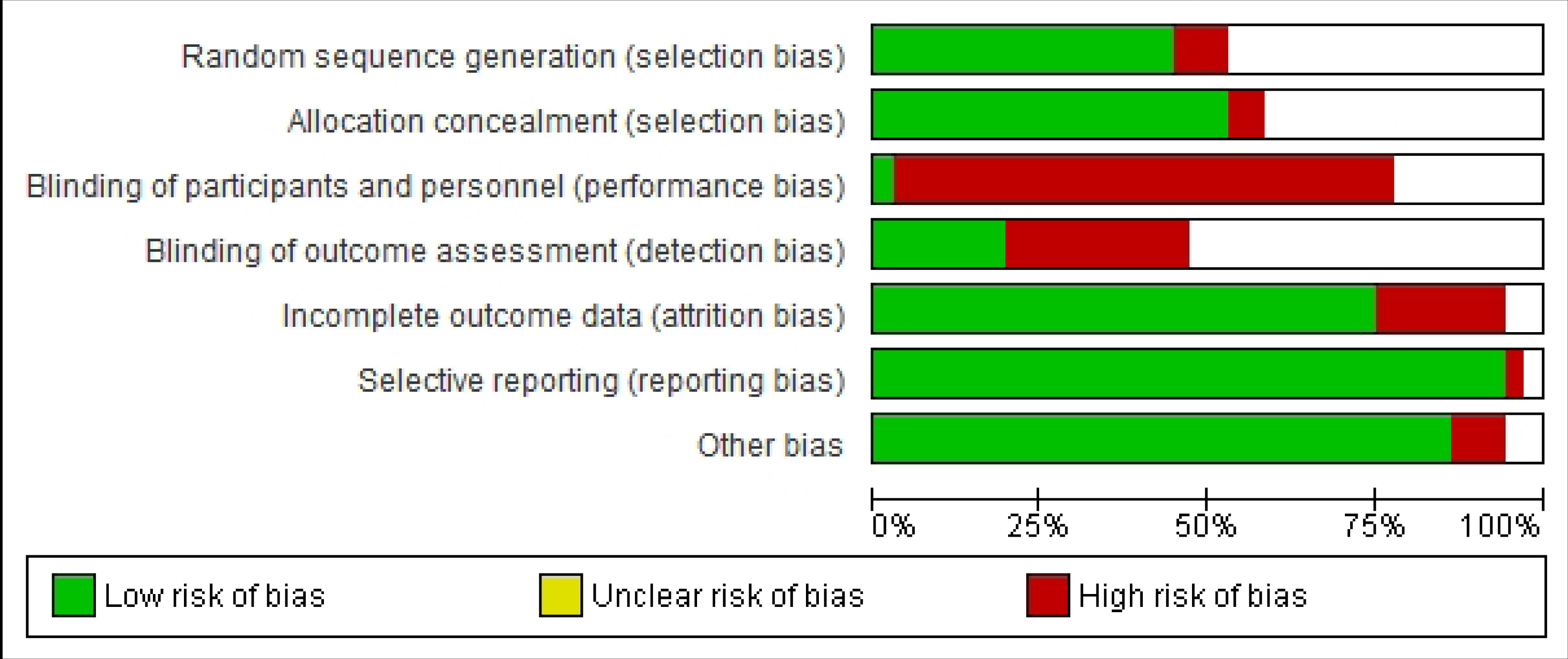
Risk of bias graph.

### Primary analysis

Table 2 reports a meta-analysis of 8 outcomes as risk ratios and standard mean differences with 95% confidence intervals. The highest overall effect is the improvement reported in the 5 min APGAR score < 7 by 1.52(95% CI 1.05,2.20). All the outcomes are statistically significant, but most show a moderate to low effect size. Considerable heterogeneity is also reported in all results except for 5-minute APGAR < 7 and instrumental delivery. Figure 4 displays the forest plot for the meta-analysis of the 5-minute APGAR score.

**Figure 4.**
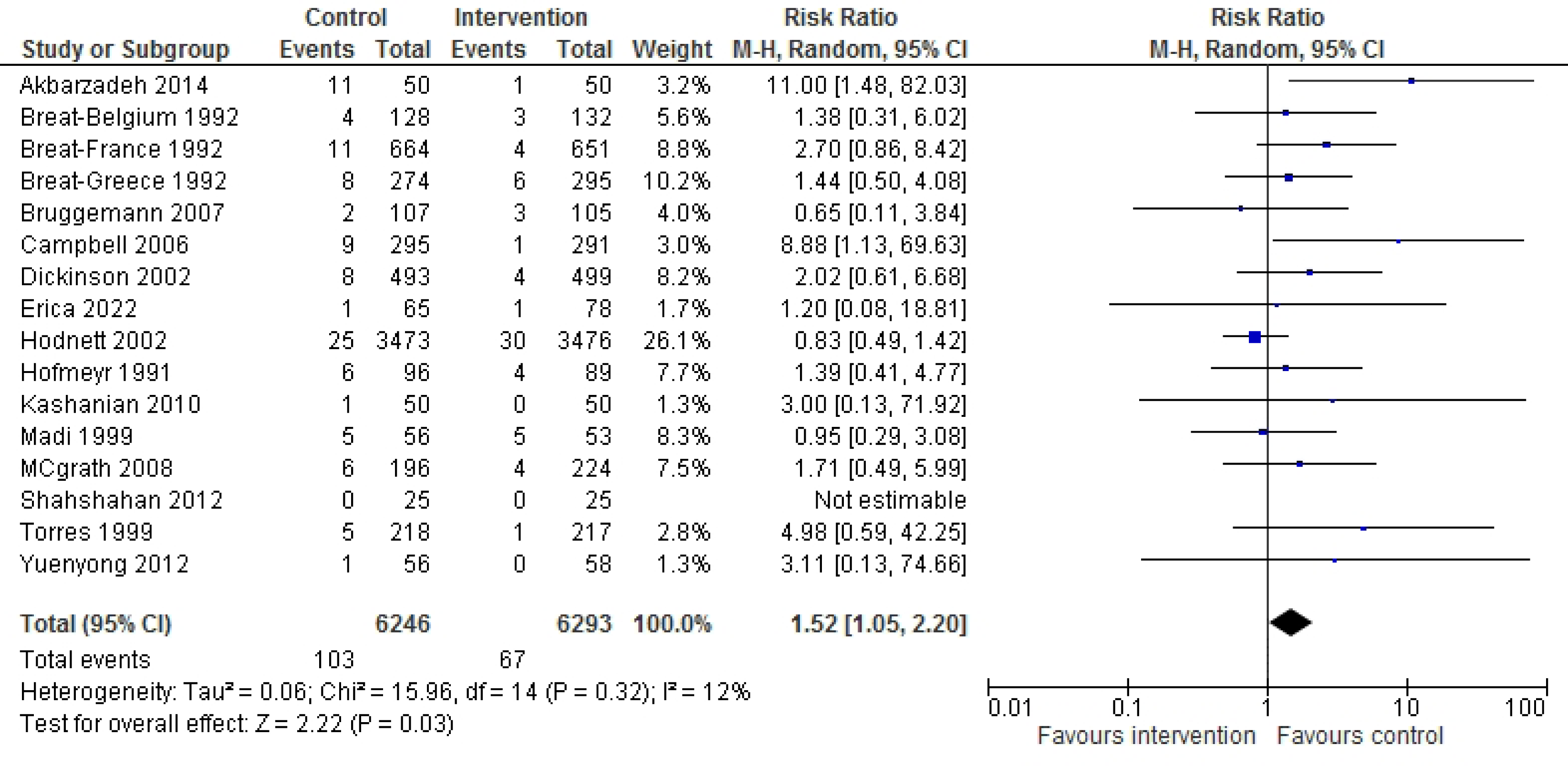
Forest plot for the meta-analysis of 5-minute APGAR score < 7.

**Table 2.** Effectiveness of a labour companion related to 8 outcomes.

| Outcome | No. of Participants (Studies) | RR (95% CI) | P value | Heterogeneity (I<sup>2</sup>) |
| --- | --- | --- | --- | --- |
| 1. Spontaneous vaginal delivery | 13811 (19 RCTs) | 1.09(1.04,1.13) | 0.0001 | 0.64 |
| 2. Duration of labour hr (Standard mean difference) | 5422 (17 RCTs) | 0.30(0.18,0.41) | 0.0001 | 0.72 |
| 3. Cesarean section | 15080 (24 RCTs) | 1.43(1.20,1.71) | 0.0001 | 0.63 |
| 4. Instrumental delivery | 13955 (21 RCTs) | 1.13(1.03,1.23) | 0.008 | 0.23 |
| 5. Oxytocin for labour induction | 12958 (21 RCTs) | 1.10(1.01,1.19) | 0.03 | 0.71 |
| 6. Analgesic usage | 12719 (18 RCTs) | 1.06(1.01,1.11) | 0.02 | 0.52 |
| 7. Tocophobia | 11133 (11 RCTs) | 1.46(1.26,1.68) | 0.0001 | 0.63 |
| 8. 5 min APGAR < 7 | 12539 (16 RCTs) | 1.52(1.05,2.20) | 0.03 | 0.12 |

### Secondary analysis

#### Familiar vs. unfamiliar labour companion

Table 3 shows the subgroup analysis of the same eight outcomes shown in Table 2 as familiar vs. unfamiliar labour companions. Among all the outcomes studied, a statistically significant subgroup difference was observed only in Tocophobia (p = 0.02), indicating that having a familiar labour companion reduces Tocophobia significantly. There was no significant subgroup heterogeneity concerning tocophobia within either subgroup. The combined effect size for Tocophobia was 1.73 (95% CI 1.49,2.02) for the familiar labour companion subgroup and 1.34 (95% CI 1.14,1.58) for the unfamiliar labour companion subgroup. However, there was an unequal distribution of trials and participants between the familiar and unfamiliar companion subgroups in all subgroup analyses. Nevertheless, for all eight outcomes, the pooled effect size for each subgroup favoured having a continuous labour companion. Figure 5 displays the forest plot for the subgroup analysis of tocophobia.

**Figure 5.**
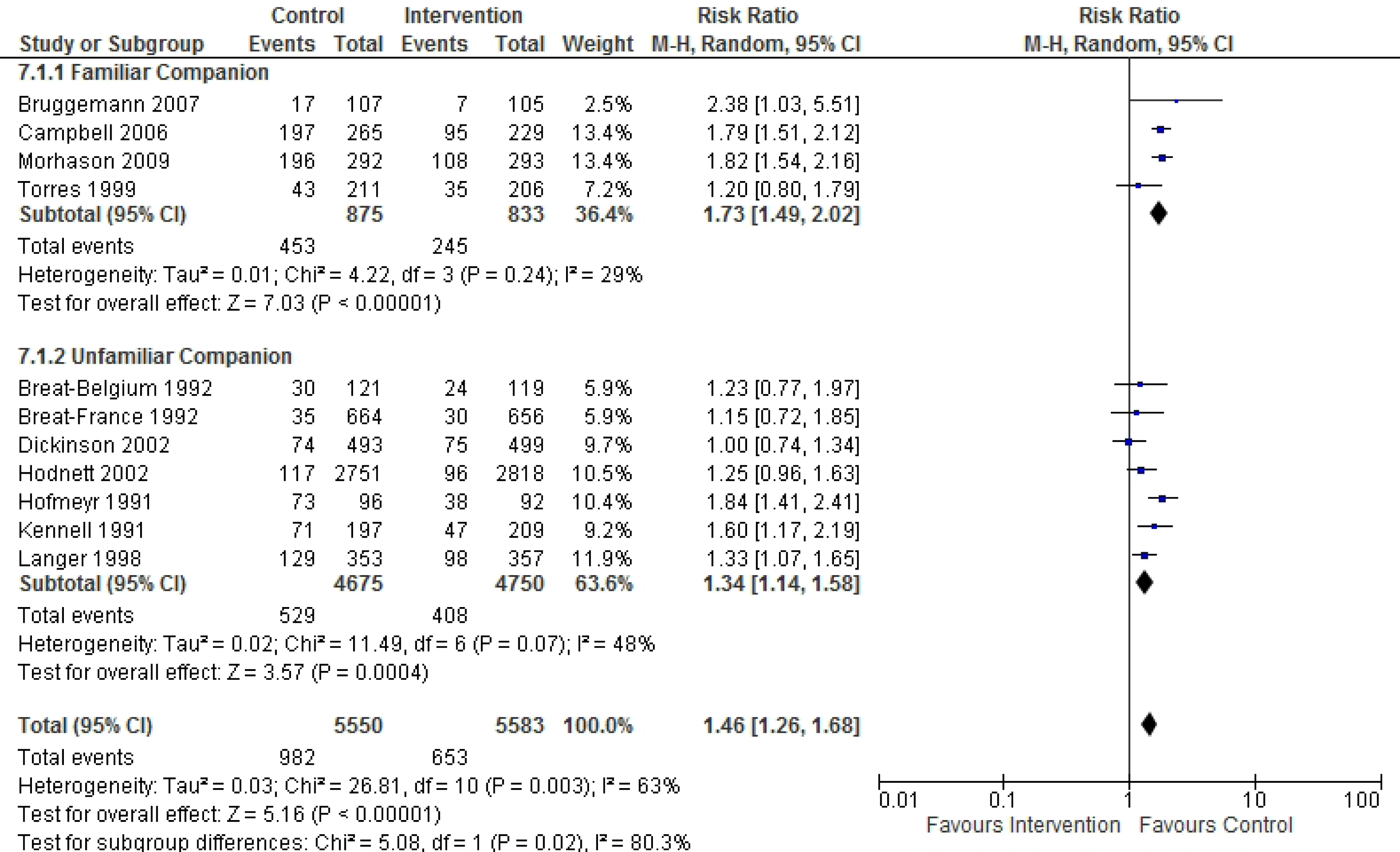
Forest plot for the subgroup analysis of tocophobia.

**Table 3:** Effectiveness of having a familiar vs. unfamiliar labour companion.

| Outcome |  | No. of Participants (Studies) | RR (95% CI) | P value | Heterogeneity (I<sup>2</sup>) | Test for subgroup difference (p) |
| --- | --- | --- | --- | --- | --- | --- |
| 1. Spontaneous vaginal delivery | Familiar | 1706 (7 RCTs) | 1.12(0.99,1.28) | 0.07 | 0.8 | 0.6 |
|  | Unfamiliar | 12105 (12 RCTs) | 1.08(1.04,1.13) | 0.0002 | 0.6 |  |
| 2. Duration of labour | Familiar | 1335 (4 RCTs) | 0.31(0.20,0.41) | 0.0001 | 0 | 0.94 |
|  | Unfamiliar | 4087 (13 RCTs) | 0.31(0.16,0.46) | 0.0001 | 0.77 |  |
| 3. Cesarean section | Familiar | 2284 (8 RCTs) | 1.55(1.00,2.39) | 0.05 | 0.76 | 0.63 |
|  | Unfamiliar | 12796 (16 RCTs) | 1.38(1.14,1.67) | 0.001 | 0.54 |  |
| 4. Instrumental delivery | Familiar | 1752 (8 RCTs) | 1.12(0.95,1.32) | 0.19 | 0.22 | 0.81 |
|  | Unfamiliar | 12203 (13 RCTs) | 1.15(1.01,1.30) | 0.03 | 0.31 |  |
| 5. Oxytocin for labour induction | Familiar | 2289 (8 RCTs) | 1.09(0.93,1.27) | 0.28 | 0.79 | 0.39 |
|  | Unfamiliar | 10669 (13 RCTs) | 1.20(1.03,1.38) | 0.02 | 0.78 |  |
| 6. Analgesic usage | Familiar | 2075 (7 RCTs) | 1.07(0.97,1.18) | 0.21 | 0.61 | 0.95 |
|  | Unfamiliar | 10644 (11 RCTs) | 1.07(1.01,1.14) | 0.03 | 0.48 |  |
| 7. Tocophobia | Familiar | 1708 (4 RCTs) | 1.73(1.49,2.02) | 0.0001 | 0.29 | 0.02 |
|  | Unfamiliar | 9425 (7 RCTs) | 1.34(1.14,1.58) | 0.0004 | 0.48 |  |
| 8. 5 min APGAR < 7 | Familiar | 1749 (8 RCTs) | 2.40(0.93,6.17) | 0.07 | 0.36 | 0.21 |
|  | Unfamiliar | 10790 (8 RCTs) | 1.25(0.88,1.79) | 0.22 | 0 |  |

#### Trained vs. untrained labour companion

Out of the analysed outcomes, only two showed statistically significant differences between having a trained labour companion versus having an untrained one: Tocophobia (p = 0.004) and the cesarean section (LSCS) rate (p = 0.02). These findings suggest that a trained companion significantly impacts these outcomes. Notably, subgroup heterogeneity concerning tocophobia was significant within the trained companion group (I2=0.59) but not in the untrained companion group (I2=0). Conversely, subgroup heterogeneity was insignificant within the trained companion group (I^2^=0.44) but significant within the untrained companion group (I^2^=0.54) in relation to the LSCS rate. For the analysis of Tocophobia, the pooled effect sizes were denoted as 1.34 (95% CI 1.14,1.57) and 1.84 (95% CI 1.60,2.12) in the trained versus untrained subgroup comparisons. At the same time, for the LSCS rate, they were represented as 1.22 (95% CI 1.05,1.42) and 2.16 (95% CI 1.37,3.40), respectively. There was a notable imbalance in the distribution of trials and participants between the trained and untrained companion subgroups across all eight outcomes. Nevertheless, in all these subgroup analyses, the pooled effect size consistently favoured the presence of a continuous labour companion. (S1 Table)

#### Effectiveness of labour companion before and after 2000

Out of the eight outcomes analysed, a statistically significant subgroup difference was observed only in the duration of labour (p = 0.004), suggesting that the classification of RCTs as before and after 2000 has a significant impact. Notably, subgroup heterogeneity was significant within the subgroup of RCTs conducted after 2000 (I^2^=0.74) but not in the subgroup of RCTs conducted before 2000 (I^2^=0.44) concerning the duration of labour. The pooled effect size for the duration of labour was 0.16 (95% CI 0.06,0.26) for the subgroup of RCTs conducted before 2000 and 0.53 (95% CI 0.30,0.77) for the subgroup of RCTs conducted after 2000. However, there was an uneven distribution of trials and participants between subgroups in all subgroup analyses. Nevertheless, having a continuous labour companion consistently showed a favorable pooled effect size in all eight outcomes. (S2 Table)

#### Effectiveness of labour companion in different geographical regions

Table 4 summarises the effect of having a labour companion in different geographical regions such as Asia, Africa, and Europe. A significant subgroup difference (p<0.05) was found in relation to the duration of labour, LSCS rate, oxytocin for labour induction, analgesic usage, and tocophobia, indicating that ethnic differences significantly influence these outcomes. The most significant overall effects are seen in Asia, followed by Africa. Effects are minimal in the European region. However, there was an uneven distribution of trials and participants between subgroups in all analyses, with the highest number of participants in the European subgroup.

**Table 4.**
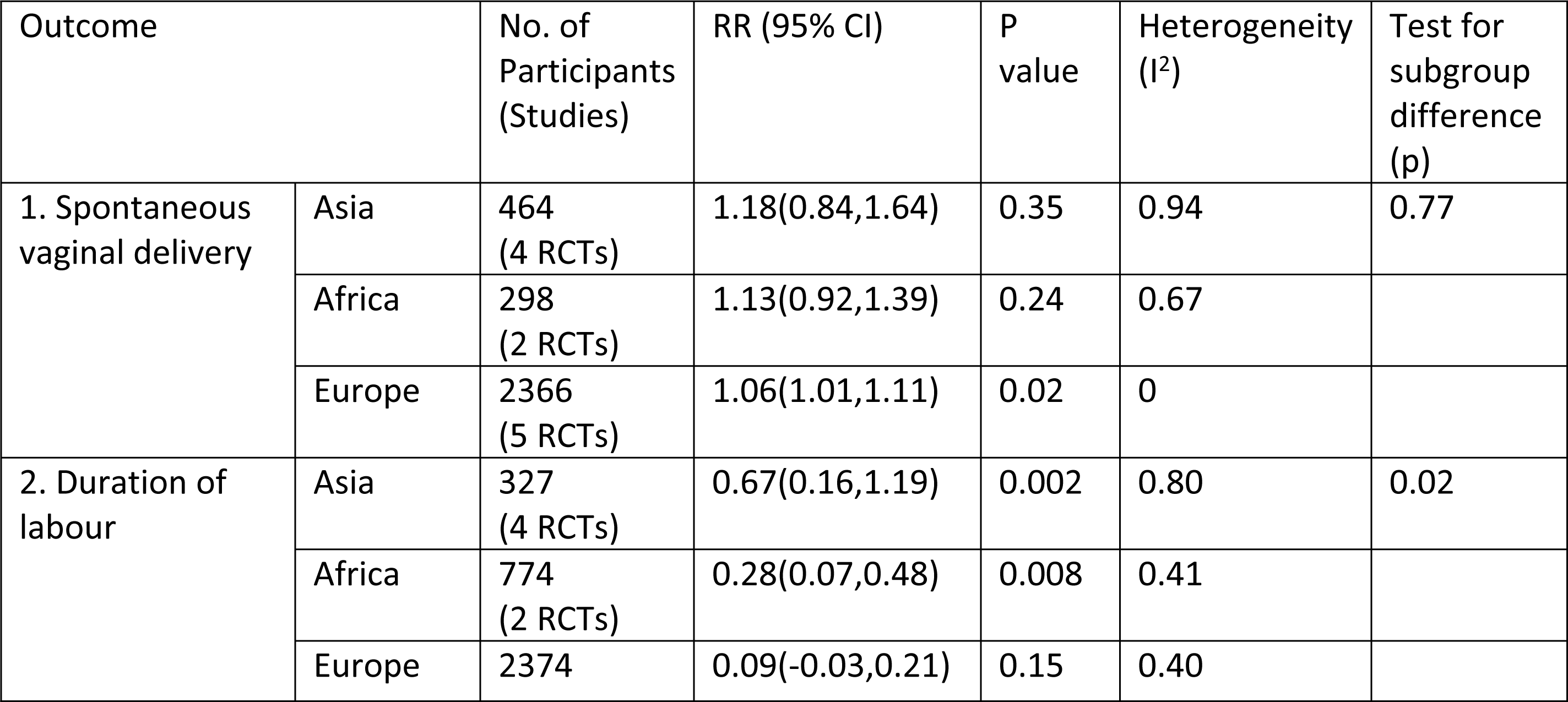
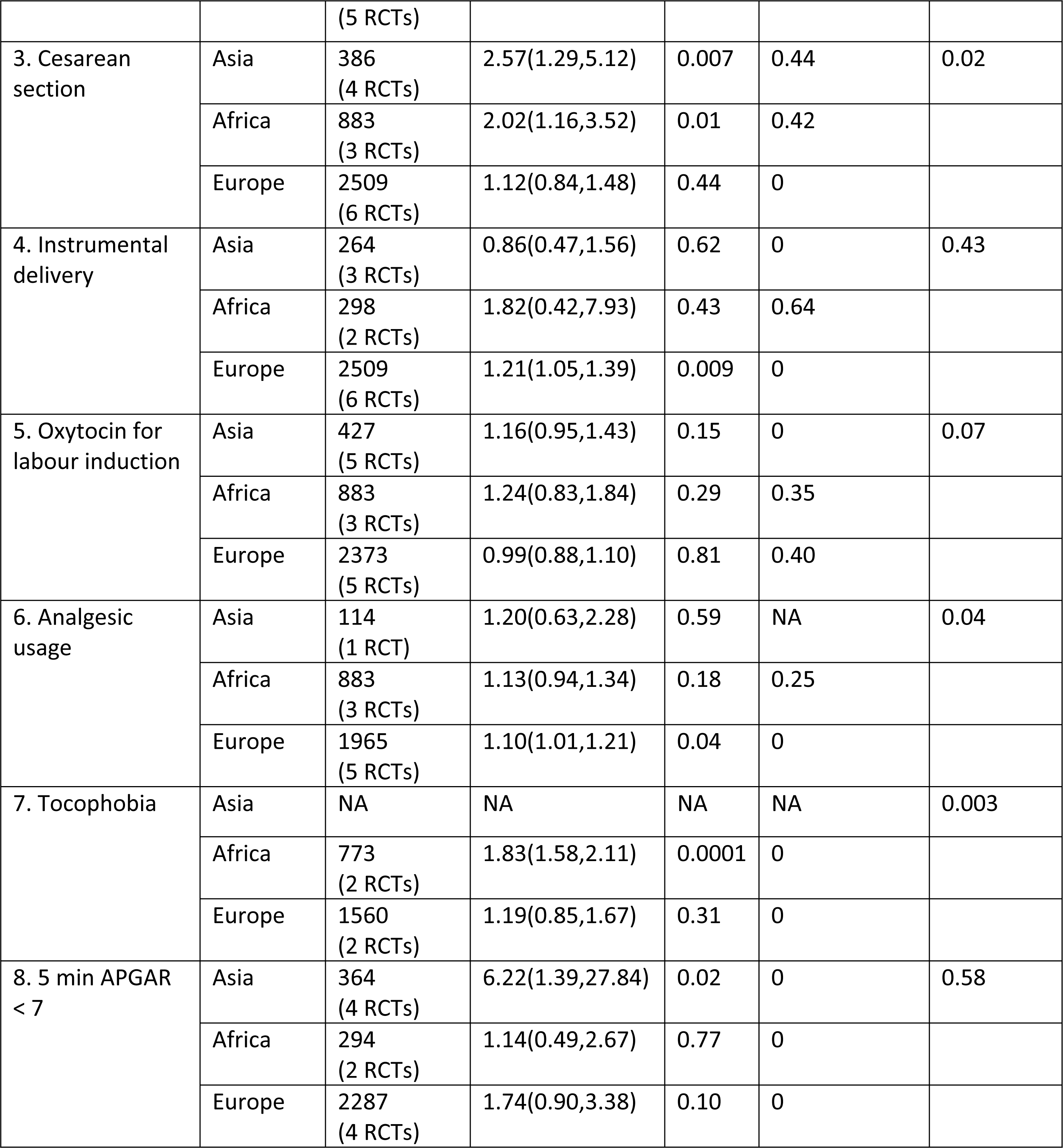
Effects of having a labour companion in different geographical regions.

## Discussion

When compared to the absence, the presence of a labour companion appears to be advantageous in all eight outcomes. All the analysed RCTs, especially the three with the highest weight, had spontaneous onset of labour as an inclusion criterion. The need to exclude labour inductions in otherwise uncomplicated pregnancies in this study group is unclear. Since spontaneous labour leads to more successful vaginal deliveries, less use of oxytocin augmentation, less assisted vaginal deliveries, shorter labours and less analgesia use compared to induced labour, the effect of a labour companion alone may not be so profound in this analysis. Studies comparing similar outcomes between spontaneous and induced labour in the presence of a labour companion would be helpful to clarify the issue.

Cesarean section rates continued to rise through the last few decades, resulting in significant medicalising of labour and resulting in higher healthcare costs. While labour companionship has a low to moderate effect on reducing Cesarean sections, it is a useful strategy to incorporate into a broader plan.

APGAR score at 5 minutes is more explicit of intrapartum fetal condition, which is improved by having a labour companion. Therefore, providing labour companionship should be considered in interventions to reduce neonatal ICU and SCBU admissions. Authors feel that the role of the labour companion does not have to stop at the baby’s delivery and may continue through the first few months of motherhood because it would be unimaginably supportive to the new mother.

Numerous studies have investigated the beneficial effects of psychological support in preventing as well as treating labour-related anxiety. Tokophobia is a form of anxiety that, in some, may amount to post-traumatic stress. Our analysis also reaffirms this with an RR of 1.46. Tokophobia also increases the sympathetic response in the mother during labour. Maternal stress reactions are associated with fetal tachycardia and variable decelerations. The resultant CTG changes may lead to high Cesarean section rates. As the management protocols used during abnormal CTG events during the studies were unavailable, the effect could not be analysed further in the available dataset.

Comparing familiar and unfamiliar labour companions revealed a larger effect size with familiar labour companions in terms of achieving successful vaginal delivery, having shorter labour, lower section rates, lower tokophobia rates, and fewer neonates with APGAR <7 at 5 minutes. Familiarity and attachment between the labouring mother and the companion provide better support during labour. Continuous encouragement, especially by a loved one, has resulted in better labour experiences and outcomes. It is uncommon to have a familiar labour companion who is also trained. Therefore, it is more likely that the effect is due to familiarity rather than the training they have received. Authors have observed that with familiar labour companions, women are less likely to adopt optimal positions during labour which are proven to shorten labour duration and reduce assisted vaginal deliveries. They are more likely to stay in the most comfortable position, which sometimes may be counterproductive. With trained support, women tend to stay in lateral or upright positions during labour without epidurals and lateral positions with epidurals.

Further, no studies determine who would be better as a labour companion. The available studies involved friends, sisters, or aunts. No studies used a partner as the labour companion. It would be interesting to compare different relatives acting as labour companions. Due to complex relationship dynamics and cultural differences, it would be very difficult to conduct and interpret such a study. Some cultures are more reluctant to accept the male partner as the labour companion. In the current resource-poor labour room setting, it would be unethical if unable to maintain the privacy of labouring women. High heterogeneity is a limitation of this study in providing recommendations on the ideal labour companion.

The current resource-poor labour room setting does not allow direct family involvement. Due to limitations in funding and staffing, there are hardly any waiting areas for the family to stay during the labour of their loved one. The labour companion may also require support during their role, and waiting areas may just be what they need to wind down. On the flip side, we also need to consider the ramifications of such an arrangement in the context of current social norms.

While most studies use low-risk mothers to mitigate the effect on labour by medical and fetal complications, there are only a handful of studies involving mothers with complicated pregnancies. For example, pregnant mothers with heart diseases would find it beneficial in terms of cardiac status to have less pain with a familiar labour companion.

There is insufficient data to assess the similarities of labour companion training associated with the included studies. Only some papers gave short descriptions of how the training was conducted. Also, healthcare workers with different levels of training and experience were employed as labour companions in the included studies. Determining the minimum training standards necessary for someone to be an effective labour companion within the current analysis is challenging. Also, developing such training programs may not be cost-effective, considering the significant advantages of having an untrained, familiar labour companion per labouring mother with trained staff to oversee the entire process. It is clear from other studies comparing psychological and emotional responses towards labouring women that training should include a psychological component. It should highlight empathy as one of the most important characteristics for a labour companion to have. Regular validation, certification and continuous professional development for these trained labour companions would be needed.

The temporal differences between studies conducted before 2000 and later may be due to changes in labour management guidelines and the availability of fetal and maternal monitoring facilities. More spontaneous vaginal deliveries may have occurred before the year 2000, while with a modern understanding of fetal physiology and the availability of continuous electronic monitoring, interventions may have become more likely. With a linear understanding of labour durations, more cesarean sections may have occurred due to a suspected lack of progress compared to the current dynamic approach to stages of labour.

Differences in effect size between Asia, Africa, and Europe may be observed due to regional differences in obstetric practice. Asia and Africa, accounting for most of the world’s developing economies, lack some basic facilities widely available on the European continent. Obstetricians, therefore, make decisions based on the overall situation rather than following strict guidelines. In using the positive influence of labour companionship, it is not surprising that more cesarean deliveries can be prevented in Asian and African regions compared to Europe. Since the cost of a cesarean delivery is much higher compared to that of a vaginal delivery, the opportunity-cost saving would be much higher.

With all the advantages, it is surprising that the labour companion is not a universal feature in the labour suites- at least in the developing world. Barriers to implementing the concept are unlikely to have been studied in an RCT.

There is a considerable preference variation among labouring women regarding their preferred labour companion, and the partner may not be the automatic and universal choice. A system that pressures the partner to accompany labour may not be evidence-based or optimal.

Our meta-analysis helps by providing the current best scientific evidence to choose and direct future exploration into the topic.

Creating awareness among the public would be essential in incorporating this paradigm-shifting practice into routine obstetric care. It would circumvent any possible backlash from different mindsets and social backgrounds.

### Strengths and limitations

We harnessed the strength of rigorous research, utilising data from 35 RCTs, and delved deeply into the nuances of labour companionship, scrutinising factors like familiarity versus unfamiliarity, trained versus untrained companions, and temporal associations - an approach not previously explored in existing meta-analyses. This comprehensive examination provides valuable insights into the impacts of labour companionship that go beyond what has been previously studied.

However, our study does come with its limitations. All 35 RCTs included in our analysis exhibited a high degree of bias, which may introduce potential sources of error. Additionally, due to significant heterogeneity in reporting across the trials, we could only identify eight distinct outcomes with sufficient studies, limiting our ability to perform robust subgroup analyses.

### Recommendations

Future research endeavours should prioritise conducting well-designed RCTs with a rigorous methodology to mitigate bias and improve the quality of evidence in this area. Additionally, efforts should be made to standardise reporting practices to enhance comparability across studies and facilitate more extensive subgroup analyses. In the interim, healthcare providers should share decision-making with expectant mothers, considering their preferences and individual circumstances when using labour companions.

## Conclusion

In conclusion, labour companionship demonstrates potential benefits across various maternal and neonatal outcomes. While it may not have as profound an effect in spontaneous labour scenarios, it still contributes to improved birthing experiences and reduced Cesarean section rates. The presence of a familiar companion appears to offer greater support during labour, emphasising the importance of emotional connection. However, there are limitations, including study heterogeneity, a lack of data on companion training, and temporal differences in study outcomes. Despite these limitations, labour companionship can be a valuable strategy to incorporate into broader obstetric care plans. Further research, standardisation of companion training, and efforts to assess acceptability and create public awareness are recommended to enhance the integration of labour companionship into routine obstetric care, potentially improving outcomes and patient experiences in labour and childbirth.

## Data Availability

All relevant data are within the manuscript and its Supporting Information files.

## Acknowledgements

None

## Supporting information

S1 Table. Effectiveness of trained vs. untrained labour companion.

S2 Table. Effectiveness of a labour companion before and after 2000.

S1 File. Analysis of spontaneous vaginal delivery.

S2 File. Analysis of duration of labour.

S3 File. Analysis of cesarean section.

S4 File. Analysis of instrumental delivery.

S5 File. Analysis of oxytocin for labour induction.

S6 File. Analysis of analgesic usage.

S7 File. Analysis of tocophobia.

S8 File. Analysis of 5 min APGAR < 7.

S9 File. PRISMA 2020 checklist.

S10 File. Detailed search strategy.

S11 File. Study protocol.

